# Spectrum of Adverse Event Following COVID-19 Immunization in High Altitude, Nepal

**DOI:** 10.1101/2022.05.19.22275337

**Authors:** Minani Gurung, Tashi Lama, Bibek Rajbhandari, Olita Shilpakar, Ashmita Maharjan, Sujita Nepal, Dev Gajmer, Kailash Lama, Lisasha Poudel

## Abstract

**Introduction:** Nepal started vaccinating frontline workers against COVID-19, in January 2021.Side effects of the vaccine are still unknown in high altitude regions. Poor surveillance and the lack of health workers in remote places to take care of people in case of vaccination side-effects could prove to be a challenge in the drive, especially in high altitudes like Humla district, Nepal. High altitude regions are harder to access and also mobilization of vaccines and manpower is strenuous due to harsh weather conditions and complex geography. We aimed to study the spectrum of Adverse Event Following COVID-19 Immunization among the front liners of Humla district.

**Methods:** This is a cross-sectional study conducted in Humla district, Nepal. COVID-19 Immunization vaccine recipients were contacted through a phone call within 5 days of vaccination to record the adverse effects. Patterns and distribution of adverse effects were analyzed in high altitude settings. Ethical approval was taken from Nepal Health Research Council.

**Results:** Of total respondents, 84.1% (95% C.I: 80.9 to 86.9) had shown symptoms after vaccination. The average time for the appearance of symptoms was 1.27 +_ 0.60 days. For systemic effects, tenderness was the side effect seen after vaccination (63.8%) followed by other side effects like pain (58.5%), Pyrexia (37.4%), Chills (29.8%), Myalgia (28.9%) and Malaise (15.2%) while for localized effects, the symptoms such as Arthralgia (16.6%) and Bruising (16.1%) were the most common effects.

**Conclusions:** Serious and life threatening adverse effects following immunization were not seen in our study site which was of altitude 2500 meter and above. Our study shows a similar type of side effect pattern as that of the lower altitude regions.

## INTRODUCTION

On January 27, 2021, Nepal’s vaccination campaign began, and as of April 20, 2021, 17 million doses of COVID vaccine had been given to chosen communities as a first dose [1]. The effectiveness of this vaccine in reducing infection, severe disease, hospitalization, and death with COVID-19 has been reported, revealing it to be 62.1% effective after two standard two doses [2]. Vaccination has altered the course of the pandemic. COVID-19 mortality and hospitalizations would have been higher without the vaccine, according to estimates.

Even though vaccines have shown excellent safety and efficacy, there have been reports of side effects. According to data submitted to the European Medicines Agency (EMA), almost 50 % of participants experienced injection site pain, headaches, or weariness after receiving the Oxford-AstraZeneca vaccine. Following the first dosage of ChAdOx1 nCoV-19 vaccine, a research done in the United Kingdom utilizing the COVID Symptom Study app found that the incidence of local and systemic reactions was 58.7% and 33.7 percent, respectively [3]. Another study employing the Mobile Vaccine Adverse Events Reporting System (MVAERS) among Korean health workers found an AEFI of 66.1 % after the first dose of ChAdOx1 nCoV-19 vaccine [4]. Shrestha et al. found an 85.04% incidence of AEFI after the first dose of Covishield immunization in one of central Nepal’s main tertiary hospitals [5].

Studies suggest that high-altitude residence may be beneficial in the novel coronavirus disease (COVID-19) implicating that traveling to high places or using hypoxic conditioning could be favorable as well [6], however there have been no studies if there were more or less side effects in high altitude.

The lack of human resources, technology to transport the vaccine, harsh climate, arduous geography is among the many challenges in the high altitude of Nepal [7]. One of the most important distribution aspects is maintaining the vaccine cold chain. Once a vial is opened, ten or eleven people must be ready to get their dose within a six-hour interval [6]. People who wish to be vaccinated have to travel by foot for many hours and sometimes even days to reach the health facilities providing the vaccine. In the same way vaccines dispatched from the capital to high altitude regions of Nepal also have a strenuous journey.

Are the side effects experienced by vaccines in high-altitude similar to other different environments? In remote, high altitudes of Nepal, many factors have to be considered and cannot be generalized with other parts of Nepal. Vaccination in higher altitude may have incompatible adverse side effects compared to the side effects of vaccination in other regions. The challenges of geography, improper cold chain maintenance, inaccessibility to better health infrastructure and the cold climate might cause contrasting effects of the vaccine. There is lack of sufficient evidence of the adverse effects of COVID19 in high altitude regions. Studying the effects of this new vaccine especially in divergent areas and understudied regions would shed light on the vaccine and add great knowledge to the literature. In this context, we conducted the study to see the spectrum of Adverse Event Following COVID-19 Immunization among frontline workers in High altitude, Humla District.

## MATERIALS AND METHODS

We conducted a telephone based cross-sectional study among 603 frontline workers between February to March 2021 to determine the spectrum of Adverse Event Following COVID-19 Immunization among frontline workers in High altitude, Humla District. Prior to data collection, ethical approval was obtained from Nepal Health Research Council (Ref no.2475). We obtained written permission from the District Administration Office in Humla and verbal consent from each participant prior to the interview. We assured voluntary participation of the participants and maintained confidentiality and anonymity of the participants throughout the study.

### Study area and study population

Our study area was Humla District. Humla is Nepal’s highest district, with most settlements located between 3,000 and 5,000 meters above sea level. Humla District, located in Karnali Province, is one of Nepal’s seventy-seven districts. The region is one of the most underdeveloped areas in Nepal with only one hospital and campus and few higher secondary schools in Simikot serving the whole district. Most of the villages of Humla don’t have access to electricity to maintain cold chain. According to the 2011 census, the district, which includes Simikot as its district headquarters, spans an area of 5,655 km2 (2,183 sq mi) and has a population of 50,858 people. The district consists of 7 Municipalities, all of which are rural municipalities. COVID-19 vaccination program in Humla district was started in February 2021 at the district hospital, Simikot. First phase of vaccination was given to frontline workers.

The target population was frontline workers who took the first dose of COVISHIELD from the district hospital located at Simikot, Humla. Participants were contacted between the 5th day of vaccination. Subjects with flu like symptoms, pregnant, lactating women, below 18 years were excluded. The first vaccine made available by the Nepalese government was COVISHIELD. This vaccine contains genetically modified organisms produced in genetically modified human embryonic kidney 293 cells.

### Sample Size calculation and sampling technique

The sample size (n) was determined using Cochrane formula, assuming 50% prevalence and 5% permissible at 95% confidence interval(CI). Taking 10% of the non-response rate, our calculated sample size was 422. However, we recruited 603 participants conveniently.

### Data collection tool and Technique

The list of recipients of the vaccine were extracted from the record available in the record section of the vaccination centers from the district hospital. We conducted telephone based interviews. The survey was estimated to take around 10 minutes to complete. We constructed and developed a questionnaire to report the AEFI. The questionnaire consisted of three components, a) Socio-demographic component including age(in years), gender(male/female), ethnicity(Brahmin, Chhetri, Madhesi, Janajati, Tharu and Dalit), education and monthly income b)anthropometric components consisting of height(in meter), weight (in kilogram) and BMI (kg/m2), and c) AEFI component (Tenderness, Pyrexia, Pain, Nausea, Myalgia, Malaise, Induration, Headache, Fatigue, Dizziness and Chills)

### Data Analysis

We analyzed the typical symptoms that have been reported. We categorized the symptoms into systemic and localized. Those diseases that affect the entire body rather than the single organ were categorized as systemic diseases whereas those which start in one area of the body or organ system and are limited to that area were categorized as localized diseases [8]. Similarly, after collecting the data, we entered, coded and analyzed data in IBM Statistical Package for Social Sciences (SPSS) version 20. We calculated frequencies and percentages for all categorical variables as a part of descriptive analysis.

## RESULTS

The mean age of respondents was 34.5 years (34.5±10.3) and the youngest respondent was of 18 years while the eldest was of 77 years. Majority of respondents (79.6%) were male. This study reported that more than a half of respondents (58.7%) belonged to the Chhetri ethnic group. Data on educational status demonstrated the average completed years of education as 11±7.1. Similarly, the average monthly income of the participants was 31443.3±18536.8.

Majority of respondents (64.5%) at the time of the study had normal BMI while very few proportion of participants had obesity. Similarly, only 12.8% of participants had one of the comorbidities.

**Table 1.**
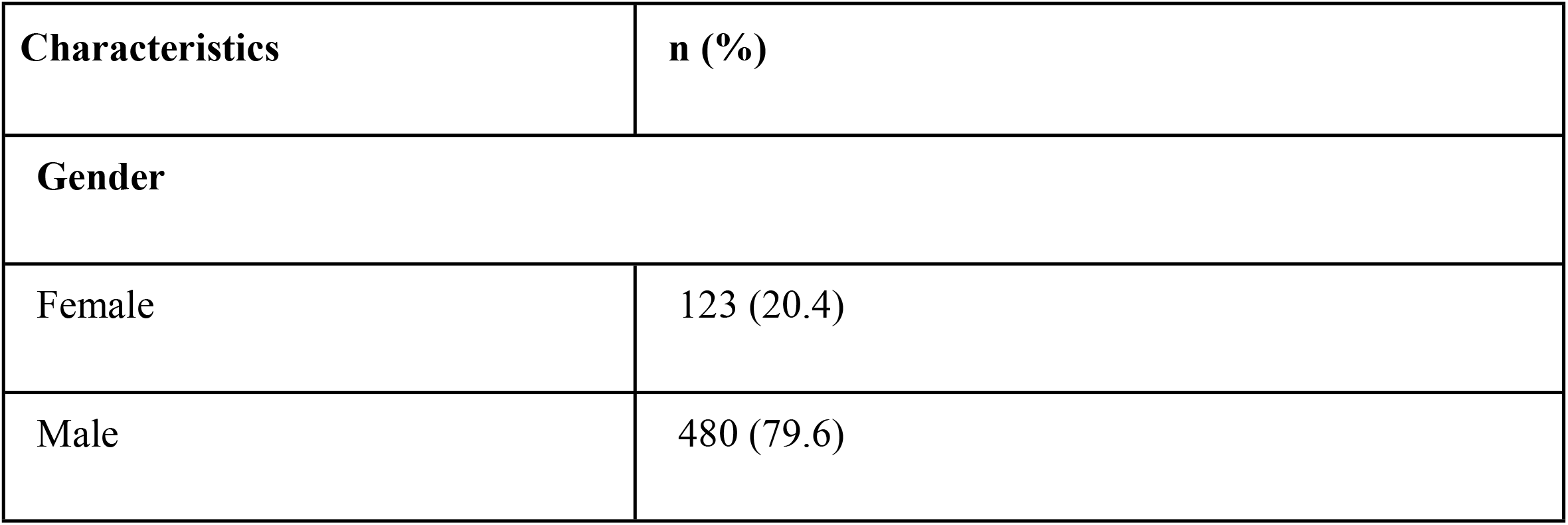
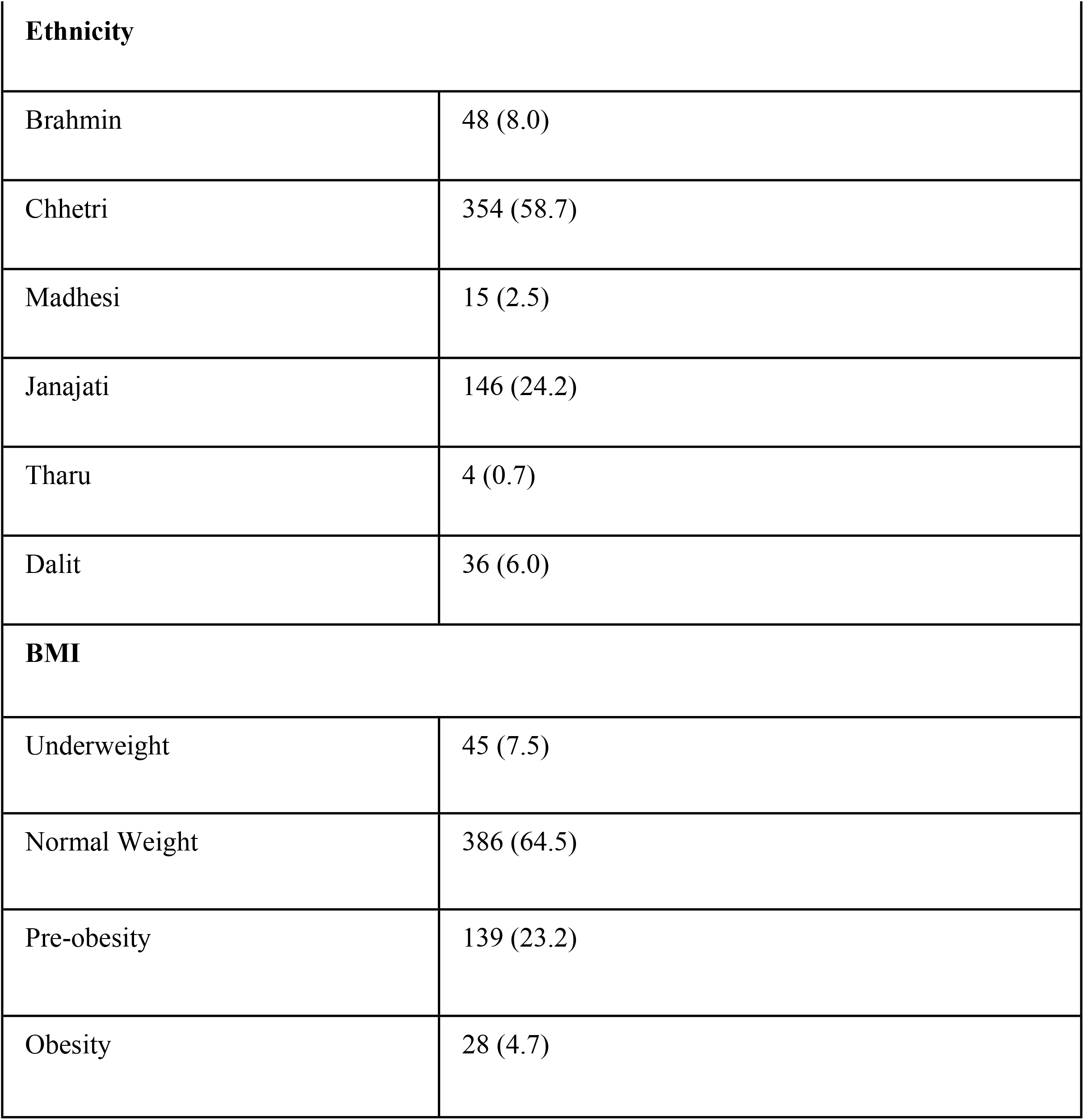
Participants‵ Characteristics (n= 603)

Of total respondents, 84.1% (95% C.I: 80.9 to 86.9) had shown symptoms after vaccination as represented by figure 1. The average time for the appearance of symptoms was 1.27 +_ 0.60 days.

**Figure 1:**
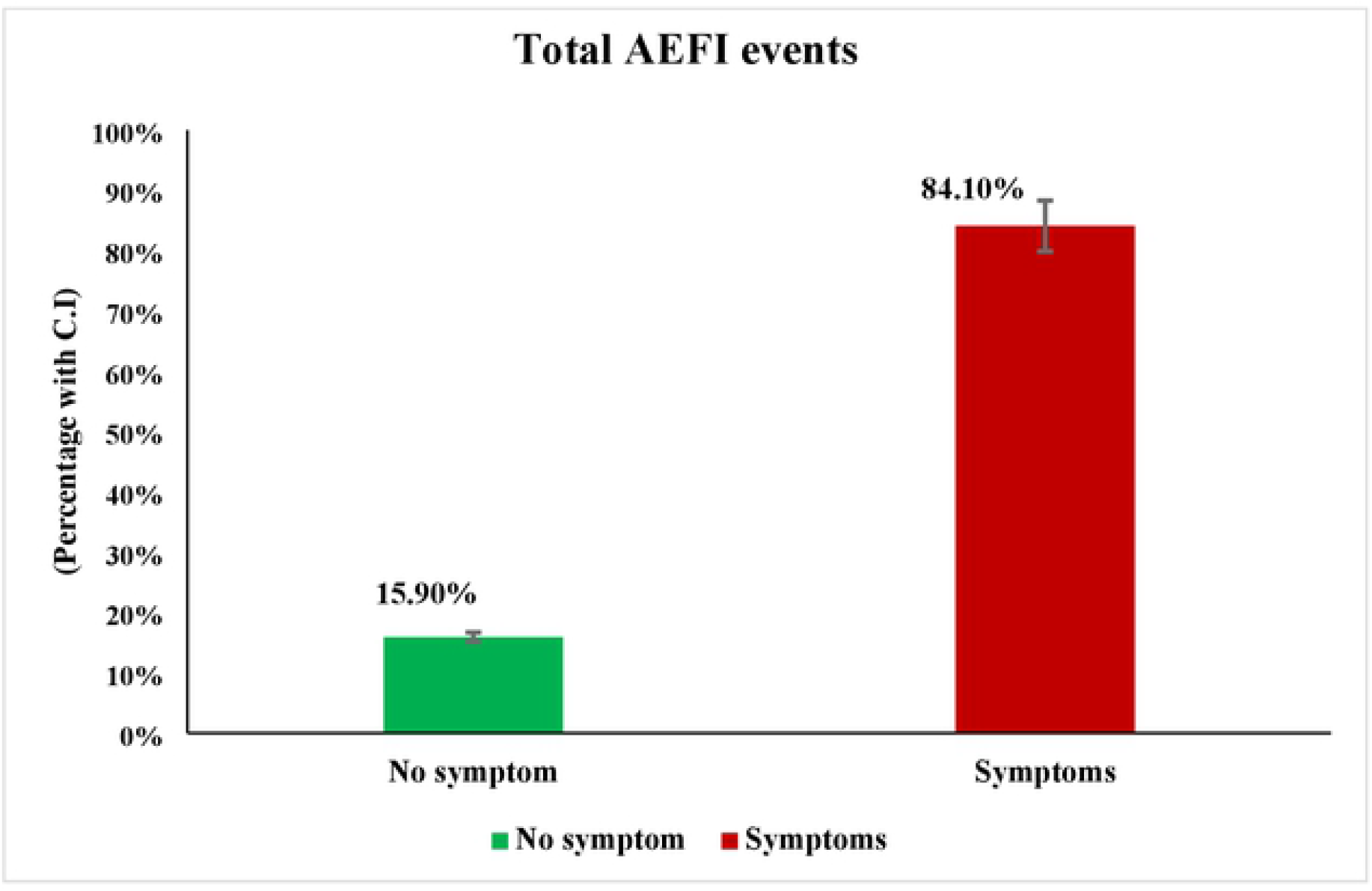
Total AEFI Events.

Figure 2 depicts the AEFI comparison between men and women, where we discovered a significant difference in the AEFI between the men and women groups.

**Figure 2:**
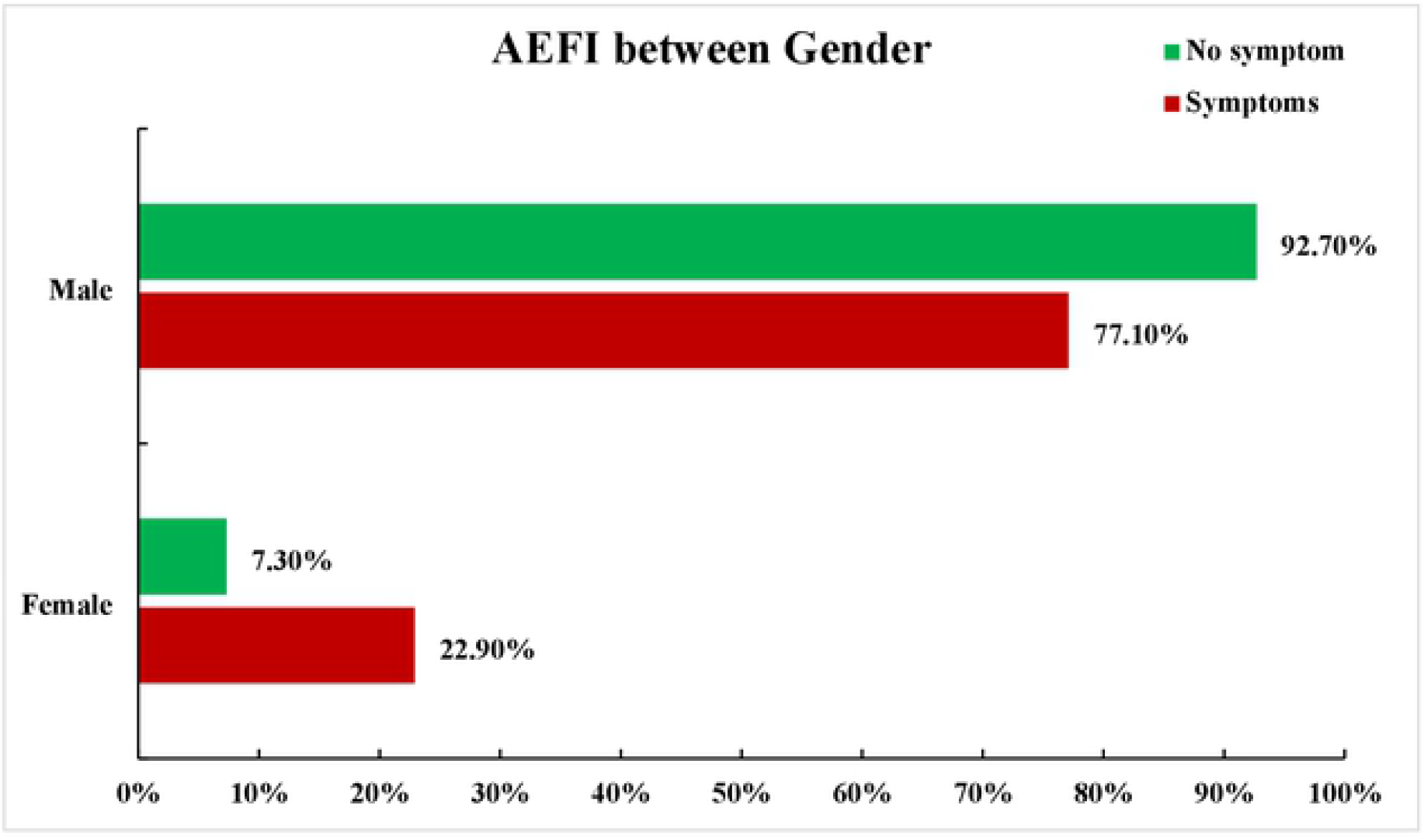
AEFI between gender.

We categorized symptoms of vaccination as systemic and localized effects. As represented in the figure 3 tenderness was the significant side effect seen after vaccination (63.8%) followed by other side effects like pain (58.5%), Pyrexia (37.4%), Chills (29.8%), Myalgia (28.9%) and Malaise (15.2%).Likewise, other effects like Dizziness, Nausea and Injection site induration were less common among the participants.

**Figure 3:**
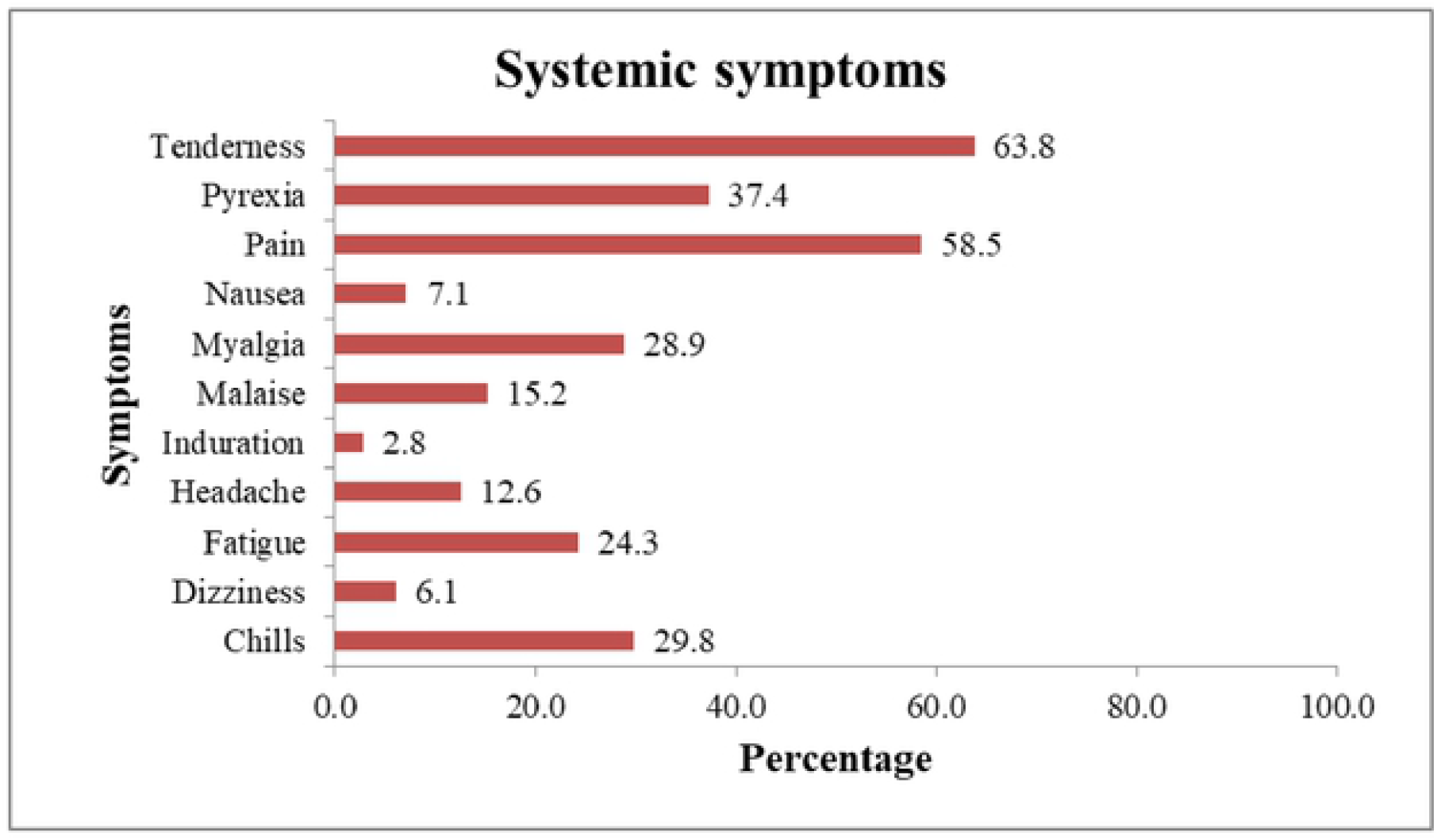
Systemic Symptoms.

**Figure 4:**
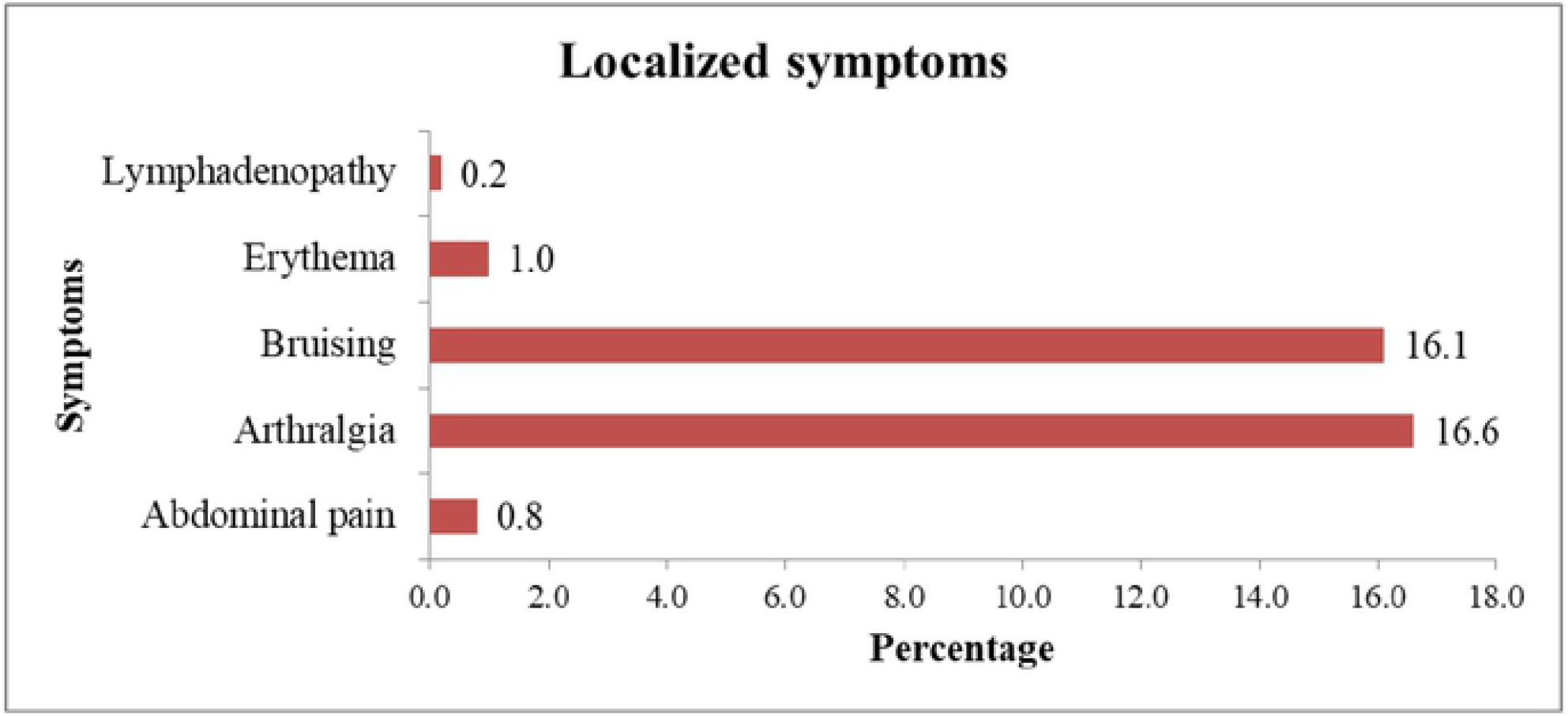
Localized symptoms.

While for localized effects the symptoms such as Arthralgia (16.6%) and Bruising (16.1%) were the most common effects followed by Erythema, Abdominal pain and Lymphadenopathy.

## DISCUSSION

This study aimed to study the spectrum of Adverse Event Following COVID-19 Immunization among the front liners in High Altitude, Humla district, Nepal. The most common side effects seen in this study were tenderness (63.8%), pain (58.5%), pyrexia (37.4%) followed by chills (29.8%), myalgia (28.9%) and fatigue (24.3%) which are common side effects of COVID-19 according to WHO and Centers for Disease Control and Prevention (CDC) [9]. This study mainly focused on AEFI in high altitude. Out of 603 respondents, the majority (84.1%) had shown AEFI events which allied with the post vaccine cross-sectional study conducted among health care workers in Chitwan Medical College of Nepal which is located in Terai region accounting for 78.9% among 424 respondents. Therefore, the percentage of adverse effects seen among participants are almost similar regardless of difference in regions [10].

Similarly, a study done in western Nepal revealed that 91.6% of the total respondents developed some form of AEFI where the majority of them were males with mean age 33 which aligns with the findings presented by our study[11]. While, one of the studies which was conducted in India showed a contrasting result as a major proportion of women reported post-vaccination symptoms in comparison to men[12]. This could be as, when compared to men, women are regarded to have a more robust immune response and build larger cell-mediated and humoral immune responses in response to antigenic stimulation by vaccination or infection [13]. Likewise, a study conducted in Jember, Indonesia did not show any association between AEFI events and gender which is opposed to the result revealed by our study [14]. This difference might be due to the dissimilarity in study population. Initially, countries like Peru, temporarily suspended the trials of Covid vaccine after finding neurological problems in one volunteer [15]. Furthermore, studies conducted in countries like Canada and Russia reported symptoms more among the female population than in male which is in contrast to the findings of our study [16,17].

This study found the majority of side effects were injection site related effects like tenderness (63.8%), pain (58.5%) and pyrexia (37.4%) which allied with most of the immunization cases [18]. Comparably, one of the studies from Japan revealed 82% of local adverse events like headache, fatigue, nausea, 71% of injection site pain and 48% of systemic adverse events like myalgia which is common in this study [19]. A study conducted in the United Kingdom in the peak winter season from December 2020 to March 2021 reported that tenderness and local pain around the site of injection were the most common symptom experienced by the participants which is consistent with the findings revealed in this study. This resemblance in both the studies might be due to similar temperate conditions [20].

## CONCLUSION

Serious and life-threatening adverse effects following immunization were not seen in our study site which was of altitude 2500 meter and above. Our study shows a similar type of side effect pattern as that of the lower altitude regions.

## Data Availability

Dataset are available in the submission file

## Data availability statement

All data are fully available without restriction.

## REFERENCES

1. Basnyat B. Nepal begins Covid-19 vaccination drive. Nepali Times. 2021 Jan 27. Available from: https://www.nepalitimes.com/banner/nepal-begins-covid-19-vaccination-drive/

2. Hung IF, Poland GA. Single-dose Oxford–AstraZeneca COVID-19 Vaccine Followed by a 12-Week Booster. The Lancet. 2021 Mar 6;397(10277):854–5.

3. Menni C, Klaser K, May A, Polidori L, Capdevila J, Louca P, Sudre CH, Nguyen LH, Drew DA, Merino J, Hu C. Vaccine Side-Effects and SARS-CoV-2 Infection After Vaccination in Users of the COVID Symptom Study App in the UK: A Prospective Observational Study. The Lancet Infectious Diseases. 2021 Jul 1;21(7):939–49.

4. Jeon M, Oh CE, Lee J. Adverse Events Following Immunization Associated with Coronavirus Disease 2019 Vaccination Reported in the Mobile Vaccine Adverse Events Reporting System. 2021 May 03;36(17).

5. Shrestha S, Devbhandari RP, Shrestha A, Aryal S, Rajbhandari P, Shakya B, Pandey P, Shrestha RK, Gupta M, Regmi A. Adverse events following the first dose of ChAdOx1 nCoV-19 (COVISHIELD) vaccine in the first phase of vaccine roll out in Nepal. Journal of Patan Academy of Health Sciences. 2021 May 15;8(1):9–17.

6. Millet GP, Debevec T, Brocherie F, Burtscher M, Burtscher J. Altitude and COVID-19: Friend or foe? A narrative review. Physiol Rep. 2021;8(24):e14615.

7. World Health Organization. Climate Change and Human Health. 2003. Available from: http://apps.who.int/iris/bitstream/handle/10665/42742/924156248X_eng.pdf;jsessionid=C9E63D25B1A7102A619232125C6D146C?sequence=1

8. The live better team. Systemic diseases versus localized diseases. 2016 Aug 29. Available from: https://reverehealth.com/live-better/systemic-diseases-versus-localized-diseases/

9. Centers for Disease Control and Prevention. Possible Side Effects After Getting a COVID-19 Vaccine. 2022. Available from: https://www.cdc.gov/coronavirus/2019-ncov/vaccines/expect/after.html

10. Adhikari P, Adhikari K, Gauli B, Sitaula D. Acceptance of COVID-19 Vaccine and Pattern of Side Effects in Nepalese Context: A Post-Vaccine Cross-Sectional Study Among Healthcare Workers in a Tertiary Care Hospital. JCMC. 2021 Jun. 19;11(2):34–8.

11. Gautam A, Dangol N, Bhattarai U, Paudel S, Poudel B, Gautam S, et al. ChAdOx1 nCoV-19 Vaccine and its Self-Reported Adverse Events: A Cross-Sectional Study from Western Nepal. J Glob Heal Reports. 2021;1–7.

12. Jayadevan R, Shenoy RS, Anithadevi TS. Survey of Symptoms Following COVID-19 Vaccination in India. medRxiv. 2021 Jan 1.

13. Fink AL, Klein SL. Sex and Gender Impact Immune Responses to Vaccines Among the Elderly. Physiology (Bethesda). 2015 Nov;30(6):408–16.

14. Supangat, Sakinah, E.N., Nugraha, M.Y. et al. COVID-19 Vaccines Programs: Adverse Events Following Immunization (AEFI) among Medical Clerkship Students in Jember, Indonesia. BMC Pharmacol Toxicol 22. 2021 Oct 12.

15. Peru suspends Chinese Covid-19 vaccine trial over serious side effects. Trt world. 2020 Dec 12. Available from: https://www.trtworld.com/americas/peru-suspends-chinese-covid-19-vaccine-trial-over-serious-side-effects-42293

16. Jarynowski A, Semenov A, Kamiński M, Belik V. Mild Adverse Events of Sputnik V Vaccine in Russia: Social Media Content Analysis of Telegram via Deep Learning. J Med Internet Res. 2021 Nov 29;23(11)

17. Government of Canada. Reported side effects following COVID-19 vaccination in Canada. 2022 April 29. Available from: https://health-infobase.canada.ca/covid-19/vaccine-safety/

18. Azarpanah H, Farhadloo M, Vahidov R, Pilote L. Vaccine Hesitancy: Evidence from an Adverse Events Following Immunization Database, and the Role of Cognitive Biases. BMC Public Health. 2021 Sep 16;21(1):1686.

19. Suehiro M, Okubo S, Nakajima K, Kanda K, Hayakawa M, Oiso S, et al. Adverse events following COVID-19 virus vaccination in Japanese young population: The first cross-sectional study conducted by a questionnaire survey after the first-time-injection. MedRxiv. 2021 Jan 1.

20. Menni C, Klaser K, May A, Polidori L, Capdevila J, Louca P, et al. Vaccine Side-Effects And Sars-Cov-2 Infection After Vaccination In Users Of The Covid Symptom Study App in the UK: A Prospective Observational Study. Lancet Infect Dis. 2021 Jul 1;21(7):939–49.

